# Scalable noninvasive amplicon-based precision sequencing (SNAPseq) for genetic diagnosis and screening of β-thalassemia and sickle cell disease using a next-generation sequencing platform

**DOI:** 10.1101/2023.06.05.23290958

**Authors:** Pragya Gupta, VR Arvinden, Priya Thakur, Rahul C Bhoyer, Vinodh Saravanakumar, Narendra Varma Gottumukkala, Sangam Giri Goswami, Mehwish Nafiz, Aditya Ramdas Iyer, Harie Vignesh, Rajat Soni, Nupur Bhargava, Padma Gunda, Suman Jain, Vivek Gupta, Sridhar Sivasubbu, Vinod Scaria, Sivaprakash Ramalingam

**Affiliations:** CSIR- Institute for Genomics and Integrative Biology, Mathura Road, Sukhdev Vihar, New Delhi, India; Academy of Scientific and Innovative Research (AcSIR), Ghaziabad, India; Thalassemia and Sickle Cell Society, Rajendra Nagar, Hyderabad, India; Government Institute of Medical Sciences (GIMS), Greater Noida, India

**Keywords:** β-thalassemia, sickle cell disease, β-hemoglobinopathies, high-throughput amplicon sequencing, next-generation sequencing, molecular diagnosis

## Abstract

β-hemoglobinopathies such as β-thalassemia (BT) and Sickle cell disease (SCD) are inherited monogenic blood disorders with significant global burden. Hence, early and affordable diagnosis can alleviate morbidity and reduce mortality given the lack of effective cure. Currently, Sanger sequencing is considered to be the gold standard genetic test for BT and SCD, but it has a very low throughput requiring multiple amplicons and more sequencing reactions to cover the entire HBB gene. To address this, we have demonstrated an extraction-free single amplicon-based approach for screening the entire β-globin gene with clinical samples using **<u>S</u>**calable **<u>n</u>**oninvasive **<u>a</u>**mplicon-based **<u>p</u>**recision **<u>seq</u>**uencing (SNAPSeq) assay catalyzing with next-generation sequencing (NGS). We optimized the assay using noninvasive buccal swab samples and simple finger prick blood for direct amplification with crude lysates. SNAPseq demonstrates 100% sensitivity and 100% specificity, having a 100% agreement with Sanger sequencing. Furthermore, to facilitate seamless reporting, we have created a much simpler automated pipeline with comprehensive resources for pathogenic mutations in BT and SCD through data integration after systematic classification of variants according to ACMG & AMP guidelines. To the best of our knowledge, this is the first report of the NGS-based high throughput SNAPseq approach for the detection of both BT and SCD in a single assay with high sensitivity.

## Introduction

β-thalassemia (BT) and sickle cell disease (SCD) are mendelian monogenic red blood cell disorders and are among the most prevalent disorders worldwide. They are caused due to genetic mutations in the β-globin (HBB) gene and have an estimated incidence of 40-82 per 1,000 live births ^1, 2^. β-hemoglobinopathies are a prevalent cause of health problems across multiple groups of ethnicities. SCD results from a single amino acid change at the 6^th^ position (G**A**G to G**T**G) in the β-globin gene to a replacement of glutamic acid to valine, whereas there are more than 200 different mutations reported for BT. These are majorly point mutations or deletions present across the HBB gene. An estimated 7% of the world’s population carries a defective β-globin gene, and around 300,000 babies are born each year with severe hemoglobin abnormalities ^3^. Recent estimates suggest that this number could rise to more than 400,000 by the year 2050 ^4^. Treatment options for β-hemoglobinopathies are primarily interventional rather than providing a permanent cure. Hydroxyurea, pain management and intravenous hydration are used in SCD whereas frequent blood transfusion coupled with iron chelation therapy are the treatment options for BT. Currently, allogeneic hematopoietic stem cell transplantation (HSCT) is the only curative option available for both the above diseases but success is primarily dependent on the availability of human leukocyte antigen (HLA)-matched donor ^5^.

Although advancements in disease management and care have been made, there is still a high incidence rate, which has a profound impact on the public health system demanding significant investment in patient care. Therefore, early detection of the disease remains the realistic approach for disease prevention, decreasing the burden on the healthcare system and aiding better disease management with the ultimate goal of reducing patient mortality and morbidity rates. At present, hemoglobin electrophoresis, high-performance liquid chromatography (HPLC) and isoelectric focusing are the predominantly used diagnostic methods. In general, HPLC tests require further confirmation through Sanger sequencing-based DNA analysis^6, 7^. Currently, most commercially available diagnostic methods rely on antibody-based recognition of Hb variants, however, these tests may provide deceptive results in blood-transfused individuals. Moreover, commonly used antibody tests in newborn screening, may lead to inconclusive results as they often undergo blood transfusion due to multiple birth-related complications ^8^. Hence, subsequent validation after 90 days of transfusion is necessary for such cases ^9, 10^. Additionally, the basal level of fetal hemoglobin expression is high in the first few months of the newborn thus limiting the applicability of antibody-based diagnostic tests. Despite advancement, the genetic diagnosis of BT continues to be challenging due to the prevalence of mutations and deletion throughout the HBB gene. Protein-based diagnostic tools cannot detect BT deletions, where 619 bp deletion is common in Southeast Asian population ^11^. Hence, a reliable diagnostic method is required that can differentiate the genotype of β-hemoglobinopathies efficiently. Besides early detection of patients, carrier detection will unveil the scope of genetic counseling in reducing the frequency of the disease. World Health Organization (WHO) indexed hemoglobin testing as one of the most crucial in vitro diagnostics (IVD) tests for primary care use in developing and underdeveloped countries^12^

The classification of genetic diseases for accurate diagnosis can be provided by allele-specific detection of disease mutation. Molecular approaches to identify point mutations are mainly dependent on methods such as gold standard traditional Sanger sequencing ^13^ and real-time PCR ^14^ which are very low throughput and need multiple amplicons and several sequencing runs with different primers pairs in order to cover the entire *HBB* gene and to identify pathogenic variants. In addition, the read quality of Sanger sequencing is often not good in the first 50-80 bases where the primers bind to start amplification. Further, different combinations of compound heterozygous mutations in different ethnic groups demand the need for an integrated diagnosis system that can identify most of the mutations associated with the disease. Moreover, it is challenging to develop a molecular diagnostic test that does not necessitate nucleic acid isolation and purification because current genotyping methods based on DNA amplification use relatively high DNA purity. Thus, establishing a molecular test that analyzes the samples without DNA extraction and purification has remained elusive. The goal of the current study is to create a low-cost, low-complexity buffer system using common laboratory chemicals that enables direct PCR amplification using a straightforward process that effectively releases DNA from buccal swabs or finger prick blood.

In the last several years, next-generation sequencing (NGS) has become a cost-effective tool for the high-throughput identification of genetic variants, thereby unveiling new opportunities in the field of molecular diagnosis^15^. When compared to Sanger sequencing, NGS-based amplicon sequencing has several advantages such as higher sequencing depth with enhanced sensitivity, enhanced discovery power and higher mutation resolution, high throughput and maximum data that can be generated with the same quantity of DNA. The diagnosis of aneuploidy in embryo biopsy specimens is made possible by this technique, which has previously been neglected for use in assisted reproductive technology^16–18^. Recently, single gene mutations in embryos have also been diagnosed using NGS; nevertheless, the clinically employed techniques still call for extensive customization in the design and optimization of multiplex-PCR for the amplification of mutant sites and polymorphisms ^14, 19, 20^. Further, Hallem et al. and Haque et al. have validated that NGS in patient-based and population-based carrier screening of inherited recessive disorders and demonstrated acceptable rates of false-positive and false-negative as well as cost-effectiveness of the tool ^21, 22^. Besides, NGS has higher sequencing depth, which enables enhanced sensitivity and high mutation resolution ^23^, which is not possible in other methods.

In the current pilot study, we demonstrated a unique strategy based on the NGS approach, for the detection of virtually all HBB mutations accountable for SCD and BT. A simple non-invasive buccal swab specimen or finger prick blood has been adopted, which is economical and can be used as a direct sample matrix avoiding any DNA extraction process. For a proof-of-concept, the robustness of the SNAPseq assay in the detection of allele-specific BT and SCD genotypes from extraction-free non-invasive crude lysates of buccal swab samples with no additional PCR cleanup has been shown with patients, carriers and wild-type samples. A systematic SNAPseq data analysis pipeline was established and validated to prioritize and predict the pathogenicity of all HBB genetic mutations identified in each specimen. The present study shows that a simplified sampling procedure combined with NGS has enormous potential and clinical utility in the molecular diagnosis of genetically heterogeneous diseases such as BT. Our study demonstrated that the NGS-based SNAPseq assay, compared to conventional sequencing approaches, can identify virtually all β-globin mutations, including compound heterozygous and large deletions in a precise manner. These results suggest that SNAPseq is a precise and most efficient platform for large-scale genetic screening and clinical genotyping in subjects with SCD and BT.

## 2. Materials and Methods

### 2.1 Clinical sample collection

The study was approved by the Institutional Ethics Committee at the Government Institute of Medical Sciences (GIMS) Greater Noida and CSIR-Institute of Genomics and Integrative Biology, New Delhi. All the participants provided written informed consent for the study. Peripheral blood samples (collected in EDTA-coated tubes, BD Biosciences) or buccal swab samples or finger prick samples were collected. Buccal swab samples were collected using nylon-flocked buccal swabs (Himedia, INDIA) to swab each inner cheek at least 10-15 times and immediately placed in the lysis buffer.

### 2.2 Sample preparation

The Buccal swabs and blood spot samples were collected from study subjects and placed in a lysis buffer containing 50mM NaOH. Each sample was mixed thoroughly for 10 minutes by vortexing and incubated at 95℃ for 10 minutes. Sample tube was allowed to cool at room temperature and then the swab was removed from the sample tube. Following, 120 µL Tris-Cl (pH-8.0) was added to each sample tube for maintaining the pH.

### 2.3 Primer design

Multiple primers were designed to amplify the entire HBB gene covering a range of BT mutations. Primers were designed using the IDT OligoAnalyser tool and synthesized from Sigma-Aldrich, India. All the primers designed are listed in Table 1.

**Table 1:**
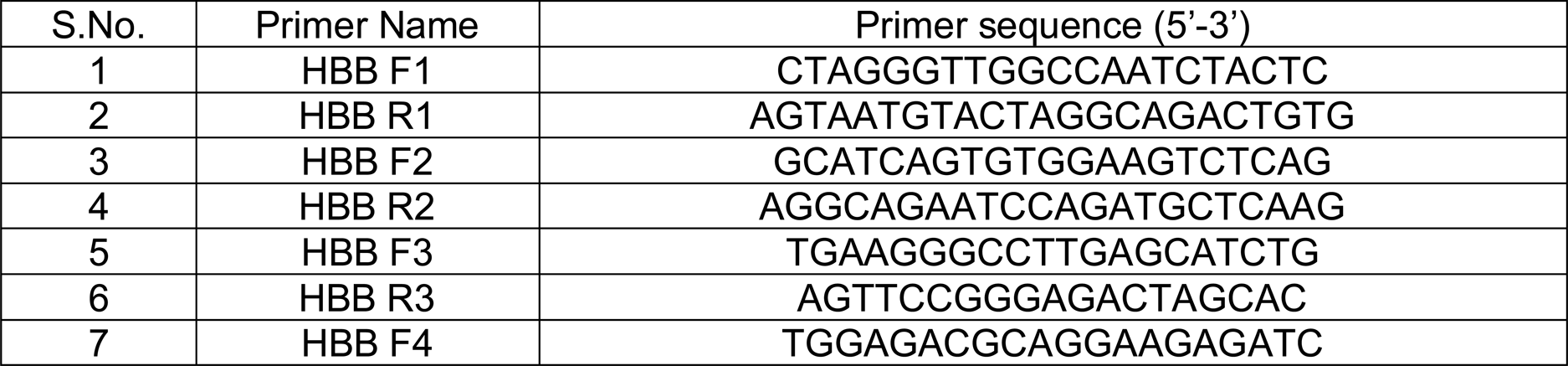
List of primers used in the study

### 2.4 Sensitivity and specificity analysis

The sensitivity of purified DNA was estimated by cloning the HBB-F4 and HBB-R3 amplicon in pJET cloning vector and was serially diluted in a range of 10^8^-10^1^ copy number. The copy number was determined by the formula: Number of copies per µL = ((Amount of ds DNA) X (6.022X10^23^ molecules per mole)) / ((length of dsDNA) X 10^9^ ng X (660 g/mole)). Each dilution was used as a template for PCR amplification using HBB-F1 and HBB-R3 primers. For buccal swab and blood samples, a range of 30% to 0.1% was used as a template in the total PCR reaction.

### 2.4 PCR amplification

PCR amplification for the entire HBB gene (includes promoter region, 5’ untranslated region (UTR), all exons, both the introns and the 3’-UTR) was carried out using various specimens collected from the patients or the volunteers (Genomic DNA, Buccal swabs and Blood) as a template. The amplification was done using PrimeSTAR max polymerase (Takara Bio Inc., Japan) in recommended concentration. The program set in thermal cycler was 1 minute at 98°C as initial denaturation, followed by 35 cycles of 15 sec at 98°C for denaturation, 15 sec at 58 °C as annealing, 15 sec at 72°C for extension, and final extension at 72°C for 5 min. The analysis of PCR amplified products was done by electrophoresis in 1% (w/v) agarose gel and visualized under Gel Documentation System (Biorad).

### PCR amplification and Sanger sequencing

Clinical samples were used for HBB gene amplification using three sets of primers to cover the entire HBB gene for Sanger sequencing. Primer combination HBB F1-HBB R1, HBB F2-HBB R2 and HBB F3 -HBB R3 was used for PCR amplification using OneTaq DNA polymerase (NEB, USA). The PCR products were analyzed by agarose gel electrophoresis and were column purified with MicroSpin columns (Genetix Biotech,India). The purified samples were subjected to Sanger sequencing using BigDye terminator v3.1 reagents (Thermo Fisher, USA) and was carried out at AgrigenomeLabs Pvt. Ltd., Kochi, India.The sequence of the primers are listed in Table 1.

### 2.4. Library preparation and sequencing

The PCR amplified products were subjected to tagmentation, where the product underwent enzymatic fragmentation followed by tagging the fragmented product with the DNA adapter sequences. Limited-cycles PCR was performed on tagmented products to add the Index 1 (i7) adapters, Index 2 (i5) adapters, and sequences required for sequencing cluster generation. The amplified library was purified using the DNA purification beads followed by the quantification using the Qubit High Sensitivity dsDNA quantification kit (Invitrogen). The quality of the final library was assessed by performing agarose gel electrophoresis (1.5%). After the quality assessment the final libraries were subjected to the paired-end sequencing on Illumina’s next-generation sequencing platform.

### 2.5. NGS data analysis

The amplicon sequencing data was analyzed with an in-house pipeline. Briefly, fastq files were trimmed using trimmomatic and the resultant fastq files were aligned to the human reference genome (GRCh38). Further, the BAM files were sorted and PCR duplicates were removed using picard MarkDuplicates. The variants were called using Varscan and annotated to RefSeq using Annovar. Finally, the variants are matched against the clinvar likely pathogenic/ pathogenic list for SCD and BT (clinvar_20230410.vcf). The entire process of data collection, data analysis and interpretation and further, the clinical reporting is envisaged to be automated with little human intervention.

## 3. Results

### 3.1 SNAPseq strategy

The design strategy adopted for the development of scalable noninvasive amplicon-based precision sequencing (SNAPseq) for genetic diagnosis and screening of β-hemoglobinopathies is shown in Fig.1. Clinical samples were collected in the forms of blood or buccal swab or finger prick. Genomic DNA was isolated from the whole blood and the other two sets of samples (buccal swab or finger prick) were treated with a lysis buffer to release the nucleic acids. PCR amplification was performed to amplify the entire HBB gene as a single amplicon. The PCR products were directly subjected to NGS library preparation as previously described. All the samples were processed in a 96-well plate. Subsequently, 96 samples were pooled together in a single tube. The pooled libraries were denatured and neutralized and a sequencing run was performed in either MiSeq or MiSeq or iSeq platform. The raw data generated in binary base call (BCL) format was analyzed using in-house developed SANPseq pipeline (Fig.1). In this study, the entire bioinformatics pipeline from data collection to clinical reporting is completely automated to reduce human intervention.

**Figure 1:**
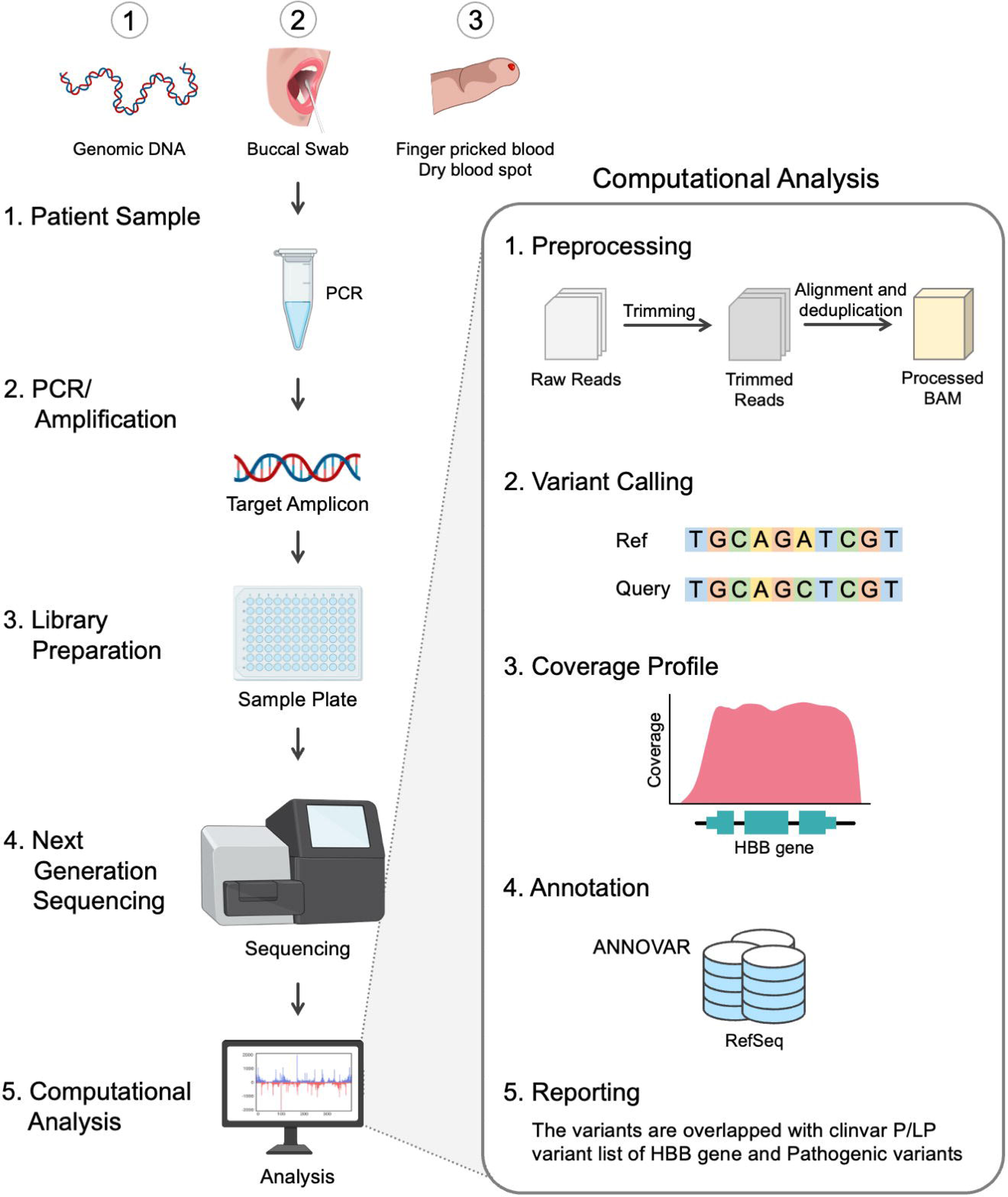
Workflow of the SNAPseq methodology

### 3.2 Standardization of primers and polymerases

The main objective of the study was to develop a high-throughput integrative assay system that can detect virtually all the mutations associated with SCD and BT in a single amplicon amplified from the HBB gene. In order to enhance the versatility of the assay, designing a primer combination that could efficiently amplify using a wider range of proofreading enzymes was required. To accomplish this, Multiple primer combinations spanning the HBB gene were designed. All primer combinations were validated for amplification using multiple proofreading enzymes including NEBNext, Phusion Polymerase, Amplitaq GOLD, Q5 polymerase, PrimeStar max polymerase, LA Taq polymerase (Supplementary Fig. 1. Our results demonstrated a consistent amplification of the HBB gene using the HBB-F1 and HBB-R3 primer combination with all the above-mentioned polymerases. The primer combination HBB-F1 and HBB-R3 covers the entire HBB gene including promoter, 5’ and 3’ untranslated regions, all three exons and both the introns of the gene (Fig. 2A, 2B). Henceforth, PrimeStar polymerase was used in the study due to its fast extension rate and high fidelity. We further investigated the reliability of NGS outcomes obtained from direct amplicons or column purified amplicons. The elimination of this step would reduce human intervention, cost and a much wider scope of automation. Hence, PCR amplification was performed using genomic DNA obtained from 8 volunteers. Our data indicate a similar median coverage 12521.45 and 12226.15 in direct amplicons and purified amplicons. Hence, these findings indicate the feasibility of direct amplicons for NGS (Fig. 2C).

**Figure 2:**
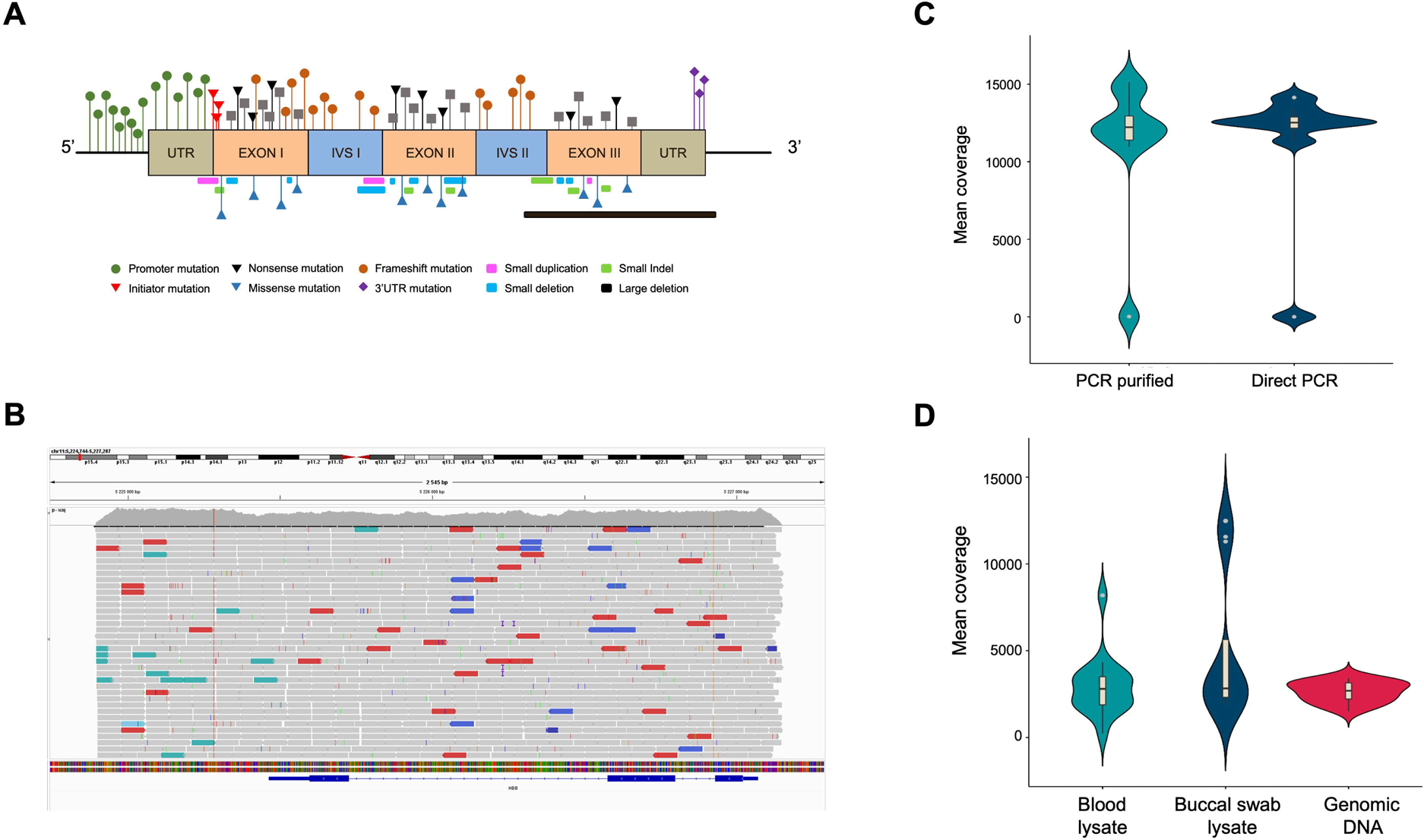
**A)** Schematics of the HBB gene highlighting all types of BT mutation spread across the gene. **B)** Tracks of the amplicon in integrative genome viewer demonstrating coverage of complete HBB gene. **C)** Violin plot demonstrating the mean coverage of PCR purified and direct PCR samples, n=10. **D)** Violin plot demonstrating the mean coverage of direct PCR samples amplified from blood lysate, buccal swab lysate and purified genomic DNA, n=16.

### 3.3 Consistent amplification with different templates

Since the process of genomic DNA isolation is arduous, time-consuming and expensive, we further investigated whether we can amplify long amplicons using direct lysate prepared from buccal swabs and finger prick blood. This approach would simplify sample collection, expedite the process and make it ideal for field deployment. Therefore, we attempted to amplify and sequence the HBB gene region from direct blood or buccal swab lysis and compared the coverage with genomic DNA. We observed a consistent amplification of the HBB gene with selected primer combinations in all three template types obtained from 8 volunteers (Supplementary Fig 2). Further, targeted sequencing resulted in similar median coverage of 2695.04, 2827.79 and 2803.20 in amplicons obtained using purified genomic DNA, buccal swab lysate and blood lysate respectively as shown in the Fig 2D. This shows the use of direct blood lysate or buccal swab lysate as template, that reproduces the similar results and hence would be useful to reduce cost and time.

### 3.4 Limit of detection

Next, we investigated the limit of detection for all three template types (purified genomic DNA, buccal swab lysate and blood lysate). In order to assess the limit of detention in purified DNA samples, a range of 10^8^-10^1^ copies/ul dilution of pHBB plasmid was used. The results suggested that polymerase is efficient enough to amplify from as low as 10 target DNA copies (Fig.3A, Supplementary Fig 3A). Direct lysates contain genomic DNA in crude form; hence it becomes challenging to calculate the copy number accurately. To address this issue, we conducted additional tests to determine the maximum percentage of direct lysate that could potentially hinder PCR reaction due to the presence of salts. Additionally, we also tested the minimum amount of lysate required for amplification. To achieve this, we performed PCR using a range of 30%-0.01% of direct lysate as a template obtained from 3 volunteers. Our results indicate that as high as 30% of direct blood lysate was not inhibitory for PCR amplification (Fig. 3B, Supplementary 3B). On the other hand, 20% of buccal swab lysates in PCR reaction was inhibitory, possibly due to the presence of fibers from buccal swab that may have been left behind during processing (Fig. 3C, Supplementary Fig. 3C). Notably, a much higher variation in amplification was observed in buccal swab lysate as compared to blood lysate due to individual collection variability. Hence, the amount of genomic DNA may vary from individual to individual due to sampling variability. Nevertheless, our data demonstrate that as low as 0.1% of both the lysates was sufficient for amplification (Fig 3B and 3C, Supplementary Fig. 3D and 3E). Overall, 1%-15% of direct lysates are sufficient to amplify enough from both lysates.

**Figure 3:**
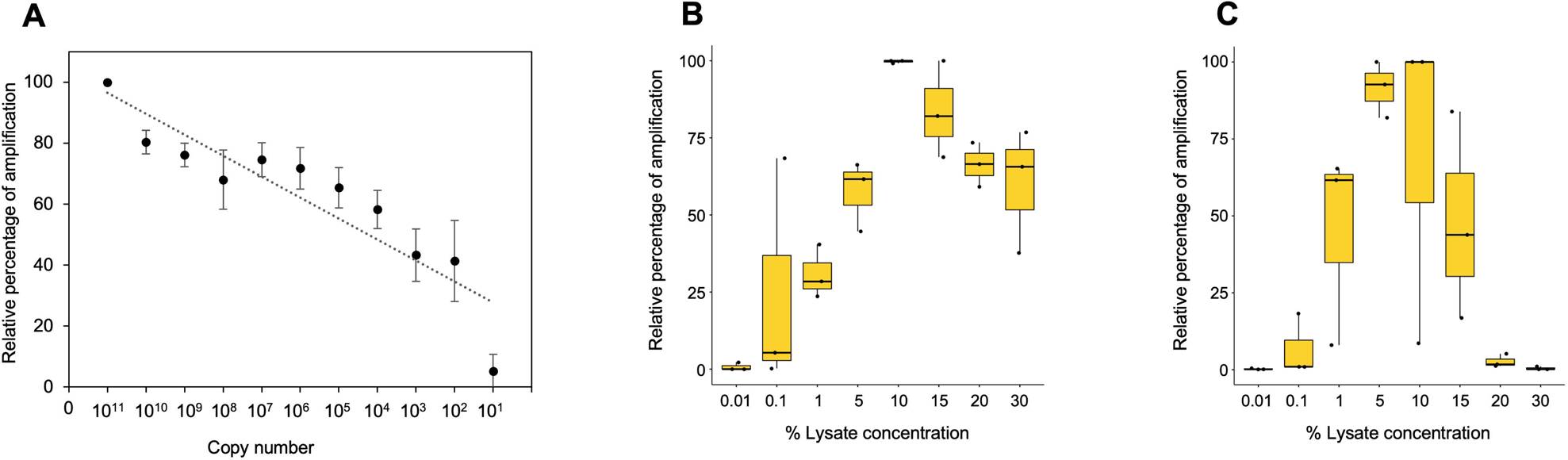
**A)** Minimum copy number required for the amplification using purified plasmid DNA. Densitometry analysis was done using ImageJ software and data from 10^11^ dilution was considered as 100%. Remaining dilutions have been plotted as percentage relative to 100% (10^11^ dilution), n=3. **B)** Box plot demonstrating minimum and maximum percentage of blood lysate required for amplification. Densitometry analysis was done using ImageJ software and the highest data value was considered as 100%. Remaining samples have been plotted as percentage relative to 100% (10^11^ dilution), n=3. **C)** Box plot demonstrating minimum and maximum percentage of buccal swab lysate required for amplification. Densitometry analysis was done using ImageJ software and the highest data value was considered as 100%. Remaining samples were plotted as percentage relative to 100% (10^11^ dilution), n=3.

### 3.5 Effect of time and temperature on crude genomic DNA lysates

As buccal swab lysates are non-invasive and can serve as a source of crude genomic DNA, we further evaluated the stability of unprocessed buccal swabs samples under variable temperatures. We have demonstrated previously that storing buccal swab samples at different temperatures (25°C, 37°C, and 42°C) prior to processing results in consistent amplification for short amplicons^24^. We adopted the same methodology to assess its impact on the amplification of large amplicons. The buccal swab samples were collected and stored at all three temperatures for 5 days followed by further processing and PCR amplification. Our results indicated consistent amplification with buccal swab sample lysates at all different temperatures (Supplementary Fig. 4A) (Table 2). To further extend the scope of sample availability, we tested if dry spots of blood can also be used as a genomic source. In a similar manner to buccal swab samples, dry spots of blood samples were stored at 25°C, 37°C, and 42°C for 5 days. Afterwards, lysates were prepared and our amplification data demonstrated consistent amplification with all the samples (Supplementary Fig.4B) (Table 2). In summary, our findings indicate that both buccal swab and blood (dry spot) lysate results in consistent amplification for NGS. We further assessed the sensitivity of sequencing by employing three different Illumina sequencing platforms: MiSeq, iSeq and MiniSeq. Our data demonstrated 2848.78, 12010.85 and 2690.19 median coverage for MiSeq, iSeq and MiniSeq respectively (Fig. 4).

**Figure 4:**
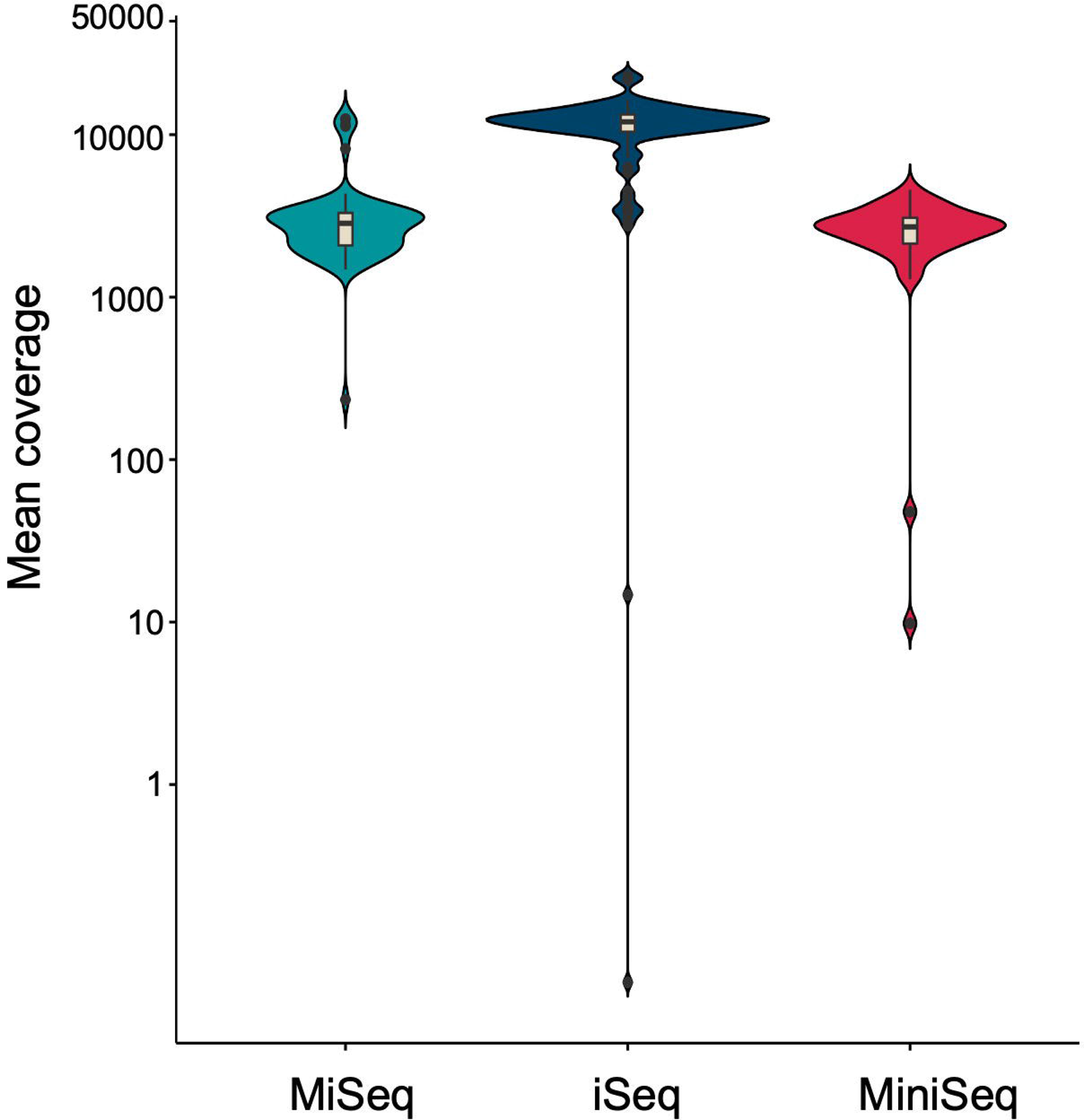
Violin plot demonstrating the mean coverage from MiSeq and iSeq sequencing platform.

**Table 2:** A comparison of PCR amplification using direct lysates stored at different temperature for 5 days

| Sample type | Storage Temperature | Number of days of storage | Sensitivity |
| --- | --- | --- | --- |
| Buccal swab | 25°C | 5 | 6/6 |
|  | 37°C | 5 | 6/6 |
|  | 42°C | 5 | 6/6 |
| Blood (dry spots) | 25°C | 5 | 5/5 |
|  | 37°C | 5 | 5/5 |
|  | 42°C | 5 | 5/5 |

### 3.6 Comprehensive and precise identification of genetic mutations associated with β-hemoglobinopathies

Next, to validate the accuracy of our pipeline for precisely identifying the genetic mutation associated with β-hemoglobinopathies (Fig. 5), we collected samples in the form of blood (for genomic DNA isolation), or buccal swab or finger prick blood. We ensured coverage of BT mutation present in the different regions of the gene as well as the compound heterozygotes including Hb S/BT, BT/BT. We amplified the target region for over 100 samples with known mutations and found that the results generated by our pipeline were in concordance with the known genotype of the sample. This indicates 100% sensitivity and 100% specificity of the SNAPseq methodology (Table 3). We further sought to test the genotype of patients clinically declared as thalassemia based on their clinical profile. We were able to identify the mutation in the patients. Further, our data indicates out of all BT patients, 25% patients were compound heterozygous for the mutations. All the different mutations from known and unknown samples identified using SNAPseq methodology for this study are listed in Table 4.

**Figure 5:**
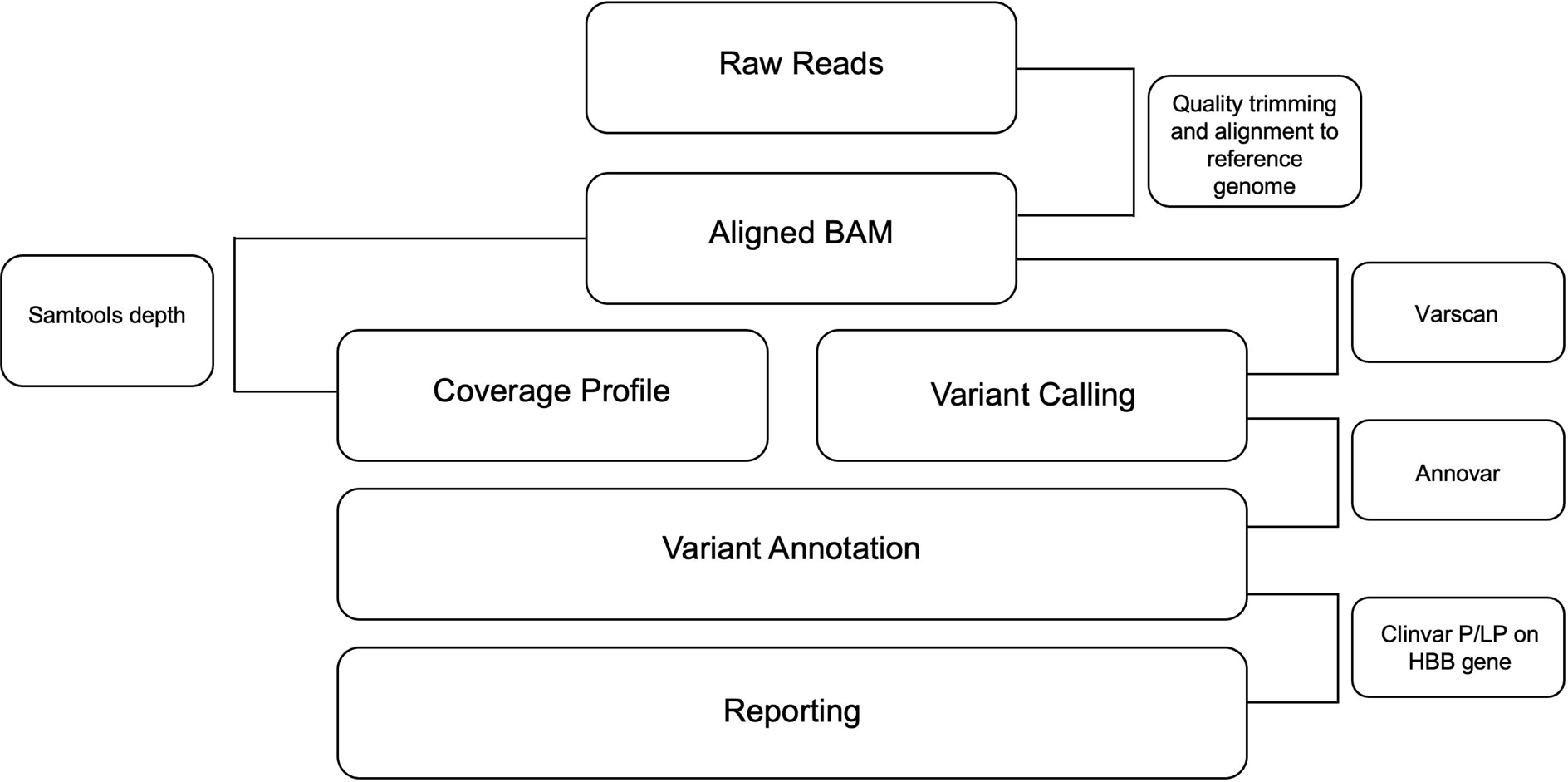
Flowchart demonstrating the pipeline for the identification of SCD and BT mutations. Sequencing data was analyzed using the inhouse pipeline. Fastq files were trimmed using the trimming algorithm and the resultant fastq files were aligned to the human reference genome. Subsequently, PCR duplicates were removed and the variants were called using Varscan and annotated to RefSeq using Annovar. Finally, the variants are matched against the clinvar likely pathogenic/ pathogenic list for SCD and BT as output from this pipeline.

**Table 3:** Sensitivity of SNAP-seq metholod

| S.No. | Genotype | Mutation Identification |
| --- | --- | --- |
| 1 | Wild Type (HbA/HbA) | 13/13 |
| 2 | Sickle cell trait (HbS/HbAA) | 10/10 |
| 3 | Sickle cell disease (HbS/HbS) | 39/39 |
| 4 | β-thalassemia | 36/36 |
| 5 | β-thalassemia heterozygous | 1/1 |
| 6 | Compound β-thalassemia | 3/3 |
| 7 | Compound β-thalassemia/<br>Sickle cell disease | 3/3 |
|  | **Total** | **105/105** |

**Table 4:** List of mutation identified in the study

| S.No. | Disease | Common name | Chromosome location | HGVS Name |
| --- | --- | --- | --- | --- |
| **β-thalassemia** |  |  |  |  |
| 1 | β-thalassemia | IVS I-5 (G>C) | Chr11-5226925 | HBB:c.92+5G>C |
| 2 | β-thalassemia | CD 8/9 (+G) | Chr11-5226994 | HBB:c.27dupG |
| 3 | β-thalassemia | CD 26 GAG>AAG | Chr11-5226943 | HBB:c.79G>A |
| 4 | β-thalassemia | CD 41/42 (-CTTT)<br>(CD 41/42 (-TTCT),<br>CD 41/42 (-TCTT)) | Chr11-5226762 | HBB:c.126_129delCTTT |
| 5 | β-thalassemia | CD 17 (+A) | Chr11-5226970 | HBB:c.53dup |
| 6 | β-thalassemia | CD 15 TGG>TAG | Chr11-5226975 | HBB:c.47G>A |
| 7 | β-thalassemia | CD 5 -CT | Chr11-5227003 | HBB:c.17_18delCT |
| 8 | β-thalassemia | IVS I-1 (G>C) | Chr11-5226930 | HBB:c.92+1G>C |
| 9 | β-thalassemia | 619 bp deletion (Asian Indian) | Chr11-5225256-5225875 | NG_000007.3:g.71609_72227del619 |
| 10 | β-thalassemia | IVS I-1 G>A | Chr11-5226930 | HBB:c.92+1G>A |
| 11 | β-thalassemia | Init CD ATG>ACG | Chr11-5227020 | HBB:c.2T>C |
| **Compound heterozogotes (β thalassemia/β-thalassemia)** |  |  |  |  |
| 1 | β-thalassemia | IVS I-5 (G>C)/<br>IVS I-1 (G>C) | Chr11-5226925/<br>Chr11-5226930 | HBB:c.92+5G>C/<br>HBB:c.92+1G>C |
| 2 | β-thalassemia | CD 26 GAG>AAG<br>/CD 15 TGG>TAG | Chr11-5226943/<br>Chr11-5226975 | HBB:c.79G>A/<br>HBB:c.47G>A |
| 3 | β-thalassemia | IVS I-5 (G>C)/<br>CD 15 TGG>TAG | Chr11-5226925/<br>Chr11-5226975 | HBB:c.92+5G>C/<br>HBB:c.47G>A |
| 4 | β-thalassemia | IVS I-5 (G>C)/<br>IVS II-1 G>A | Chr11-5226925/<br>Chr11-5226576 | HBB:c.92+5G>C/<br>HBB:c.315+1G>A |
| 5 | β-thalassemia | IVS I-5 (G>C)/CD<br>41/42 (-CTTT) (CD<br>41/42 (-TTCT), CD<br>41/42 (-TCTT)) | Chr11-5226925/<br>Chr11-5226762 | HBB:c.92+5G>C/<br>HBB:c.126_129delCTTT |
| 6 | β-thalassemia | IVS I-5 (G>C)/<br>CD 54 -T | Chr11-5226925/<br>Chr11-5226725 | HBB:c.92+5G>C/<br>HBB:c.165delT |
| 7 | β-thalassemia | IVS I-5 (G>C)/<br>IVS II-837 (T>G) | Chr11-5226925<br>Chr11-5225740 | HBB:c.92+5G>C/<br>HBB:c.316-14T>G |
| 8 | β-thalassemia | VS I-1 (G>C)/<br>CD 8/9 (+G) | Chr11-5226930 /<br>Chr11-5226994 | HBB:c.92+5G>C/<br>HBB:c.27dupG |
| **Compound heterozyotes for HbS/β-thalassemia** |  |  |  |  |
| 1 | HbS/<br>β-thalassemia | CD 6 (GAG>GTG)/<br>IVS I-5 (G>C) | Chr11-5227002/<br>Chr11-5226925 | (HBB):c.20A>T/<br>HBB:c.92+5G>C |
| 2 | HbS/<br>β-thalassemia | CD 6 (GAG>GTG)/<br>41/42 (-CTTT) (CD<br>41/42 (-TTCT), CD<br>41/42 (-TCTT)) | Chr11-5227002/<br>Chr11-5226762 | (HBB):c.20A>T/<br>HBB:c.126_129delCTTT |
| 3 | HbS/<br>β-thalassemia | CD 6 (GAG>GTG)/<br>619bp deletion | Chr11-5227002/<br>Chr11-5225256-5225875 | (HBB):c.20A>T/ |
|  |  |  |  | NG_000007.3:g.71609_72227del619 |
| Sickle cell disease (HbSS) |  |  |  |  |
| 1 | Sickle cell disease (HbS/HbS) | CD 6 (GAG>GTG) | Chr11-5227002 | (HBB):c.20A>T/<br>(HBB):c.20A>T |
| 2 | Sickle cell trait (HbS/HbA) | CD 6 (GAG>GTG)/<br>WT | Chr11-5227002 | (HBB):c.20A>T / WT |

In order to evaluate the robustness and reproducibility of the SNAPseq assay, we assessed the above protocol at a secondary health care center, where sample collection, sample processing, PCR amplification, NGS library preparation and sequencing run was performed by the laboratory technicians and good quality data was obtained. This clearly demonstrates the reproducibility and robustness of the assay and how easily it can be adopted in a healthcare set-up for high-throughput screening.

## Discussion

Hemoglobinopathies are the most common monogenic disorder and impose a significant global health burden on families and the health care system across the globe. Hence the early diagnosis of hemoglobinopathies is imperative for both prevention and appropriate treatment for patients. It is also crucial to provide counseling to couples and families who may be at risk to suffer severe hematological symptoms. Despite extensive studies at biochemical, molecular and hematological levels, the differential diagnosis of hemoglobinopathies remains challenging.

At present, the diagnosis of β-hemoglobin disorders is an extensive process that involves multiple stages of screening: evaluation of the blood panel and specialized hemoglobin tests for characterization. The majority of newborn screening (NBS) programs utilize protein-based hemoglobin separation methods such as isoelectric focusing (IEF), HPLC, and gel or liquid-based electrophoresis ^25^. Though protein-based techniques are the basis of hemoglobin diagnostics, they may not be able to identify BT mutations or to distinguish between HbSS and compound heterozygosity for HbS and hereditary persistence of fetal hemoglobin (HbS/HPFH) ^26^. Hence, additional confirmation of mutation is required by DNA sequencing. A low MCH (Mean corpuscular hemoglobin) and MCV (Mean corpuscular volume) are common symptoms in hemoglobinopathies, but these symptoms may often be misdiagnosed for vitamin B12, iron and folic acid deficiency ^27^. If β-hemoglobinopathy is suspected with the above symptoms, a series of specialized hemoglobin tests are recommended. Thus, multiple stages of diagnoses may lead to misdiagnosis, delayed treatment and missing out carriers of mutations who require counseling ^26^. In addition, the diagnostic differentiation is now time-sensitive and crucial due to the rising use of early, pre-symptomatic disease-modifying medication (e.g., initiation of hydroxyurea therapy at 6 months of age)^28^.

The rising prevalence of compound heterozygotes of BT and SCD possesses an additional challenge for point-of care or simple molecular based genetic tests. Similar to variable phenotypes associated with BT, a compound heterozygote of BT and SCD results in variable clinical presentation depending on the mutation. Around 10% to 15% of sickle cell disease is caused by a combination of the sickle cell and BT mutations, also known as compound heterozygosity^29^, which is not possible to detect in point-of-care screening methods. The distinction between HbSS and HbS/β genotypes using current hemoglobin-based diagnostic methods can be difficult ^30, 31^. As a result, there is a need for a scalable and comprehensive genetic diagnostic method that can identify virtually all HBB mutations associated with β-hemoglobinopathies.

NGS has transformed genomics research and created better sequencing methods. Although Sanger sequencing is competent for sequencing amplicons up to 1 kb ^32^, screening of longer templates needs primer walking or shotgun sequencing, which are more complicated and expensive ^33, 34^. The ability of SNAPseq to evaluate long amplicons is a significant advantage in such cases. SNAPseq also has a notable advantage over Sanger sequencing as they frequently provided full coverage for the target amplicon. Generation of entire coverage through Sanger sequencing necessitates bidirectional analysis in order to avoid the interpretational complications brought on by “dye blobs” and ambiguity in base calling at the start of each sequence, which can vary from 60 to 120 bases of the sequence^35^. Though Sanger sequencing can sequence up to 1 kb, the quality of the reads from 1 kb would be only around 700-800 bp due to the above reason.

NGS based molecular diagnosis has been successfully applied for various genomic applications ^36^. Recently, Kubikova et. Al, have used an NGS based approach for the diagnosis of β-hemoglobinopathies in preimplantation embryos ^37^. In this study, 8 pairs of primers were used to amplify 12 different PCR fragments spanning the coding regions of the HBB gene and splice acceptor and donor sites using purified genomic DNA extracted from participant samples. There are many BT mutations which are present in the intronic regions that are not completely covered in the study and amplification of multiple fragments with different primer sets using purified genomic DNA is an expensive, time consuming and cumbersome process, which are the major limitations of the above study. Another study by He et al reported a thalassemia carrier screening using NGS based approach but the study was designed to identify only the most common disease-causing point mutations or selected region ^38^. SNAPseq, an NGS-based approach is designed to identify both common and rare, annotated and novel variants in carriers with and without BT and sickle cell trait phenotypes, significantly improving the detection of carrier status and subsequently improving the detection rate of at-risk couples. This is in contrast with conventional carrier screening assays, which are designed to search for only the most prevalent mutations within a gene and coverage of less comprehensive due to various factors^21^.

Advancements in the field of NGS and its high-depth sequencing to identify mutations and variants have fast-tracked the diagnosis of many human genetic disorders and have been more reliable in characterizing the disease genotype than other molecular diagnostic tests available ^22^. Significant progress in this field in recent years has found numerous applications in research and genomic medicine. However, an extraction-free, universal single amplicon sequencing strategy for identifying virtually all HBB mutations has not been established. In this pilot study, we designed, optimized and furnished a SNAPseq methodology by utilizing the advantage of inexpensive tools, to identify all the associated mutations with SCD and BT in the HBB gene, using crude lysates of buccal swabs and/or blood and analyzed using an established pipeline to prioritize pathogenic mutations with allele-specific sensitivity. In addition, in this study, we evaluated the ability of two major Illumina sequencing platforms, MiSeq, MiniSeq and iSeq, which covers both high throughput and low throughput samples and obtained similar results. Our optimized assay pipeline system provides clinically relevant sensitivity in mutation calling across an HBB gene.

The current study focuses on developing a single amplicon-based strategy for screening of β-hemoglobinopathies. During the development of this assay system, one of the main challenges was to address the method of sample collection and processing, ensuring that the samples were collected in the quickest and most efficient way without compromising the accuracy and sensitivity of the test. Establishing a genetic test that does not require genomic DNA isolation and purification is challenging since existing molecular tests based on DNA amplification require relatively high purity of genomic DNA ^24^. Hence, developing a molecular test that analyzes the samples without genomic DNA isolation has remained evasive. Here, we sought to establish a cost-effective and low-complexity buffer system with common laboratory reagents that make direct genomic DNA amplification feasible by employing a simple protocol that enables the release of DNA most effectively. We have also compared our extraction-free protocol with PCR products amplified using purified genomic DNA of the same samples and obtained a similar outcome. In addition, the SNAPseq assay system has the additional advantage of no additional PCR cleanup steps required for the downstream applications (NGS) so that it reduces the overall assay cost. These cost reductions will have more benefits from the above steps in high throughput settings than from further advances in sequencing capacity.

Blood samples have been traditionally used for molecular studies and clinical diagnostics. Nevertheless, blood collection for genomic DNA isolation is an invasive procedure and demands skilled healthcare professionals^39^. Hence, in this study, we have demonstrated a crude lysate prepared from a simple finger prick blood, which does not necessitate professional assistance, results in consistent amplification. Furthermore, saliva is a more suitable and minimally intrusive alternative to blood for molecular diagnostics. However, some study participants have trouble spitting into a tube, while others have trouble producing enough saliva for genetic testing ^40^. In addition, saliva collection can be challenging due to the common medication side effect of dry mouth. However, buccal swab sample collection is rapid, non-invasive, and cost-effective, taking all this into account we indicated the feasibility of genotyping SCD and BT using buccal swab samples in this study. Additionally, it would make it simple for patients and families to collect their samples at their homes or clinics.

The maintenance of sample integrity is very crucial, especially in remote areas. Ideally, biological samples should be stored in temperature controlled and properly qualified storage and should be processed as quickly as possible after collection^41^. Hence, sample storage is another obstacle during sample collection since most of the collected samples need to be stored and transported before processing in a realistic scenario. Consequently, we have demonstrated that buccal swab samples in the collection buffer are stable at 25°C, 37°C, and 42°C for at least up to 5 days. In addition, our study also demonstrated that finger prick blood samples are also stable for at least up to 5 days as dry spots. Our study highlights the crucial significance of our findings for low-and middle-income nations with a high prevalence of β-hemoglobinopathies. These countries often face limited access to expensive refrigerated transport and adequate storage facilities for biological samples.

In this study, we provide a general workflow for the interpretation of the enormous amount of data produced through NGS-based molecular diagnosis. Using a SNAPseq approach, we analyzed over 250 samples with 100% sensitivity using a single amplification covering virtually all the HBB mutations with a simple, cost-effective and extraction-free platform. In addition to SCD and BT mutations, SNAseq can identify other genotypes such as hemoglobin type C genotypes AC, CC (HBC) and SC (compound heterozygous for HbC and HbS) and hemoglobin E (HBE) β-thalassemia mutation (point mutation in codon 26 GAG to AAG) on first allele and any other BT mutation in the second allele. Here, we have outlined a highly sensitive NGS-based carrier screening system that has a number of advantages over conventional genotyping techniques and exhibits excellent levels of precision, reproducibility, and resilience for clinical usage. The immense potential and clinical use of NGS-based SNAPseq is emphasized by a high diagnostic yield attained by the identification of genetic mutations of all forms of inheritance, particularly de novo mutations.

A similar diagnostic workflow can also be easily adopted for screening other genetic diseases with very less customization. It can offer a quick, simple, and potentially affordable high-throughput approach for screening genetic disease to detect a wide range of genotypes including point mutations, large deletions and indels. It provides a great opportunity for better assessment, disease management and genetic counseling.

In summary, we developed a highly sensitive comprehensive and precise molecular diagnostic tool SNAPseq with the combination of minimal work-up, high-throughput large-scale screening with systematic analysis and interpretation of all identified genetic mutations for β-hemoglobinopathies, a most common monogenic disease. It might be appropriate for large-scale screening where the prevalence of β-hemoglobinopathies is high.

## Data Availability

All data produced in the present work are contained in the manuscript

## Acknowledgement

This work was supported by the Council for Scientific and Industrial Research (CSIR) grant (MLP2001) Government of India (GoI) to SR. PT, PG, SG, AVR and NB are recipients of Senior Research Fellowship from Department of Biotechnology, Council of Scientific and Industrial Research, University Grant Commission and Indian Council for Medical Research, GoI respectively.

## Notes

**Conflict of Interest:** The authors declare that they have no conflict of interest

### Competing Interest Statement

The authors have declared no competing interest.

### Funding Statement

This work was supported by the Council for Scientific and Industrial Research (CSIR) grant Government of India (GoI) to SR. PT, PG, SG, AVR and NB are recipients of Senior Research Fellowship from Department of Biotechnology, Council of Scientific and Industrial Research, University Grant Commission and Indian Council for Medical Research, GoI respectively.

### Author Declarations

Ethics committee of IGIB and GIMS gave ethical approval for this work

## References

1. Piel FB, Hay SI, Gupta S, Weatherall DJ, Williams TN. Global burden of sickle cell anaemia in children under five, 2010-2050: modelling based on demographics, excess mortality, and interventions. PLoS Med, 2013, 10:e1001484

2. Weatherall DJ, Williams TN, Allen SJ, O’Donnell A. The population genetics and dynamics of the thalassemias. Hematol Oncol Clin North Am, 2010, 24:1021–31

3. Joint WHO-March of Dimes Meeting on Management of Birth Defects and Haemoglobin Disorders (2nd: 2006: Geneva, Switzerland), World Health Organization, March of Dimes. Management of birth defects and haemoglobin disorders: report of a joint WHO-March of Dimes meeting, Geneva, Switzerland, 17-19 May 2006. World Health Organization, 2006

4. Piel FB, Steinberg MH, Rees DC. Sickle Cell Disease. N Engl J Med, 2017, 376:1561–73

5. Ngo DA, Steinberg MH. Genomic approaches to identifying targets for treating β hemoglobinopathies. BMC Med Genomics, 2015, 8:44

6. Arishi WA, Alhadrami HA, Zourob M. Techniques for the Detection of Sickle Cell Disease: A Review. Micromachines (Basel), 2021, 12

7. Nair S. Potential pithfalls in using HPLC and its interpretation in diagnosing HbS. J Rare Dis Res Treat, 2018, 3:9–12

8. Whyte RK, Jefferies AL, Canadian Paediatric Society, Fetus and Newborn Committee. Red blood cell transfusion in newborn infants. Paediatr Child Health, 2014, 19:213–22

9. Kanter J, Telen MJ, Hoppe C, Roberts CL, Kim JS, Yang X. Validation of a novel point of care testing device for sickle cell disease. BMC Med, 2015, 13:225

10. Reed W, Lane PA, Lorey F, Bojanowski J, Glass M, Louie RR, Lubin BH, Vichinsky EP. Sickle-cell disease not identified by newborn screening because of prior transfusion. J Pediatr, 2000, 136:248–50

11. Lama R, Yusof W, Shrestha TR, Hanafi S, Bhattarai M, Hassan R, Zilfalil BA. Prevalence and Distribution of Major β-Thalassemia Mutations and HbE/β-Thalassemia Variant in Nepalese Ethnic Groups. Hematol Oncol Stem Cell Ther, 2022, 15:279–84

12. Second WHO Model List of Essential In Vitro Diagnostics, 2019. https://www.who.int/publications/i/item/WHO-MVP-EMP-2019.05. (accessed April 26, 2023)

13. Ngo HT, Gandra N, Fales AM, Taylor SM, Vo-Dinh T. Sensitive DNA detection and SNP discrimination using ultrabright SERS nanorattles and magnetic beads for malaria diagnostics. Biosens Bioelectron, 2016, 81:8–14

14. Chen Y, Sun J, Xianyu Y, Yin B, Niu Y, Wang S, Cao F, Zhang X, Wang Y, Jiang X. A dual-readout chemiluminescent-gold lateral flow test for multiplex and ultrasensitive detection of disease biomarkers in real samples. Nanoscale, 2016, 8:15205–12

15. Korf BR, Rehm HL. New approaches to molecular diagnosis. JAMA, 2013, 309:1511–21

16. Fiorentino F, Biricik A, Bono S, Spizzichino L, Cotroneo E, Cottone G, Kokocinski F, Michel C-E. Development and validation of a next-generation sequencing-based protocol for 24-chromosome aneuploidy screening of embryos. Fertil Steril, 2014, 101:1375–82

17. Wells D, Kaur K, Grifo J, Glassner M, Taylor JC, Fragouli E, Munne S. Clinical utilisation of a rapid low-pass whole genome sequencing technique for the diagnosis of aneuploidy in human embryos prior to implantation. J Med Genet, 2014, 51:553–62

18. Zheng H, Jin H, Liu L, Liu J, Wang W-H. Application of next-generation sequencing for 24-chromosome aneuploidy screening of human preimplantation embryos. Mol Cytogenet, 2015, 8:38

19. Yan L, Huang L, Xu L, Huang J, Ma F, Zhu X, Tang Y, Liu M, Lian Y, Liu P, Li R, Lu S, Tang F, Qiao J, Xie XS. Live births after simultaneous avoidance of monogenic diseases and chromosome abnormality by next-generation sequencing with linkage analyses. Proc Natl Acad Sci U S A, 2015, 112:15964–9

20. Ren Y, Zhi X, Zhu X, Huang J, Lian Y, Li R, Jin H, Zhang Y, Zhang W, Nie Y, Wei Y, Liu Z, Song D, Liu P, Qiao J, Yan L. Clinical applications of MARSALA for preimplantation genetic diagnosis of spinal muscular atrophy. J Genet Genomics, 2016, 43:541–7

21. Hallam S, Nelson H, Greger V, Perreault-Micale C, Davie J, Faulkner N, Neitzel D, Casey K, Umbarger MA, Chennagiri N, Kramer AC, Porreca GJ, Kennedy CJ. Validation for clinical use of, and initial clinical experience with, a novel approach to population-based carrier screening using high-throughput, next-generation DNA sequencing. J Mol Diagn, 2014, 16:180–9

22. Haque IS, Lazarin GA, Kang HP, Evans EA, Goldberg JD, Wapner RJ. Modeled Fetal Risk of Genetic Diseases Identified by Expanded Carrier Screening. JAMA, 2016, 316:734–42

23. Koboldt DC. Best practices for variant calling in clinical sequencing. Genome Med, 2020, 12:91

24. Thakur P, Gupta P, Bhargava N, Soni R, Varma Gottumukkala N, Goswami SG, Kharya G, Saravanakumar V, Gunda P, Jain S, Dass J, Aggarwal M, Ramalingam S. A Simple, Cost-Effective, and Extraction-Free Molecular Diagnostic Test for Sickle Cell Disease Using a Noninvasive Buccal Swab Specimen for a Limited-Resource Setting. Diagnostics (Basel), 2022, 12

25. Therrell BL Jr, Lloyd-Puryear MA, Eckman JR, Mann MY. Newborn screening for sickle cell diseases in the United States: A review of data spanning 2 decades. Semin Perinatol, 2015, 39:238–51

26. Shook LM, Haygood D, Quinn CT. Clinical Utility of the Addition of Molecular Genetic Testing to Newborn Screening for Sickle Cell Anemia. Front Med, 2021, 8:734305

27. Prueksaritanond S, Barbaryan A, Mirrakhimov AE, Liana P, Ali AM, Gilman AD. A puzzle of hemolytic anemia, iron and vitamin B12 deficiencies in a 52-year-old male. Case Rep Hematol, 2013, 2013:708489

28. Ware RE, McGann PT, Quinn CT. Hydroxyurea for children with sickle cell anemia: Prescribe it early and often. Pediatr Blood Cancer, 2019, 66:e27778

29. Aygun B, Bello A, Thompson AA, Davis L, Sun Y, Luo H-Y, Cui S, Chui DHK. Clinical phenotypes of three children with sickle cell disease caused by HbS/Sicilian (δβ) -thalassemia deletion. Am J Hematol, 2022, 97:E156–8

30. Uçucu S, Karabıyık T, Azik F. Difficulties in the diagnosis of HbS/beta thalassemia: Really a mild disease? J Med Biochem, 2022, 41:32–9

31. Chandra S, Ali M, Mishra P, Kapoor AK, Jindal Y. Detection of Compound Heterozygous Sickle Cell-β Thalassaemia in a Patient with Extreme Weakness, Mild Jaundice and Moderate Anaemia - A Case Report. J Clin Diagn Res, 2017, 11:ED07–8

32. Heather JM, Chain B. The sequence of sequencers: The history of sequencing DNA. Genomics, 2016, 107:1–8

33. McMurray AA, Sulston JE, Quail MA. Short-insert libraries as a method of problem solving in genome sequencing. Genome Res, 1998, 8:562–6

34. Emonet SF, Grard G, Brisbarre NM, Moureau GN, Temmam S, Charrel RN, de Lamballerie X. Long PCR Product Sequencing (LoPPS): a shotgun-based approach to sequence long PCR products. Nat Protoc, 2007, 2:340–6

35. Kieleczawa J. DNA Sequencing: Optimizing the Process and Analysis. Jones & Bartlett Learning, 2005

36. Taylor MK, Williams EP, Wongsurawat T, Jenjaroenpun P, Nookaew I, Jonsson CB. Amplicon-Based, Next-Generation Sequencing Approaches to Characterize Single Nucleotide Polymorphisms of Species. Front Cell Infect Microbiol, 2020, 10:565591

37. Kubikova N, Babariya D, Sarasa J, Spath K, Alfarawati S, Wells D. Clinical application of a protocol based on universal next-generation sequencing for the diagnosis of beta-thalassaemia and sickle cell anaemia in preimplantation embryos. Reprod Biomed Online, 2018, 37:136–44

38. He J, Song W, Yang J, Lu S, Yuan Y, Guo J, Zhang J, Ye K, Yang F, Long F, Peng Z, Yu H, Cheng L, Zhu B. Next-generation sequencing improves thalassemia carrier screening among premarital adults in a high prevalence population: the Dai nationality, China. Genet Med, 2017, 19:1022–31

39. Javaid MA, Ahmed AS, Durand R, Tran SD. Saliva as a diagnostic tool for oral and systemic diseases. J Oral Biol Craniofac Res, 2016, 6:66–75

40. Ang JS, Aloise MN, Dawes D, Dempster MG, Fraser R, Paterson A, Stanley P, Suarez-Gonzalez A, Dawes M, Katzov-Eckert H. Evaluation of buccal swabs for pharmacogenetics. BMC Res Notes, 2018, 11:382

41. Vaught JB, Henderson MK. Biological sample collection, processing, storage and information management. IARC Sci Publ, 2011:23–42

